# A cross sectional survey of Australian and New Zealand specialist trainees’ research experiences and outputs

**DOI:** 10.1101/2024.03.11.24303739

**Authors:** Paulina Stehlik, Caitlyn Withers, Rachel Bourke, Adrian Barnett, Caitlin Brandenburg, Christy Noble, Alexandra Bannach-Brown, Gerben Keijzers, Ian Scott, Paul Glasziou, Emma Veysey, Sharon Mickan, Mark Morgan, Hitesh Joshi, Kirsty Forrest, Thomas Campbell, David Henry

## Abstract

**Objective:** To explore medical trainees’ experiences and views concerning college-mandated research projects.

**Setting:** Online survey (Apr-Dec 2021) of current and recent past trainees of Australian and New Zealand colleges recruited through 11 principal colleges and snowballing.

**Participants:** Current trainee or completed training in the past 5 years.

**Main outcome measures:** We asked participants: whether they were required to conduct research as part of their college training, how they conducted their research, and their research activity after training. Respondents were invited to submit project reports for reporting and methodological quality evaluation. Data were analysed descriptively.

**Results:** Of the 372 respondents, 313 (86%) were required to complete one or more projects. Of the 177 who had completed their project (representing 267 projects), 76 provided information on 92 studies, with 34 reports submitted for evaluation. Most respondents developed their own research questions, study design and protocol, and conducted research in their own time, with 56% (38/68) stating they had the skills to complete their project. Most project teams consisted of their own medical specialty followed by statisticians, but seldom others.

44% (30/68) were satisfied with their research experience, and 53% (36/67) supported mandatory projects. Half (87/174) felt research was important for career development, 72% (44/61) considered initiating research post-training, and 54% (33/61) participated in it.

Commonly expressed themes were time-burden of conducting projects, production of research waste, and the importance of research for skills development. Of the 34 submitted reports, 75% were published and 82% had a clear research question. Only three had a low risk of bias.

**Conclusion:** Majority of respondents conducted projects, but few shared details or reports. Despite valuing their research experiences and seeing clinical relevance, time conflicts and research waste were common concerns. Colleges should focus on enhanced research methods training and creating trainee research collaboratives.

**Protocol registration:** https://doi.org/10.17605/OSF.IO/BNGZK

**Summary box:** Majority of medical specialty trainees are required to conduct a research project to develop their research skills.

We found the learning experiences are inconsistent, and the quality of research produced even more so.

A new approach is required that is tailored to the research skills required by most practicing clinicians, namely being expert in applying research to practice and in participating in collaborative research. Those wishing to become leaders in research should be supported to do so via a specialised well-supported pathway.

## Background

Medical specialty training colleges often require doctors to conduct research to earn their professional qualifications. Such practices are widespread, including in the UK, Europe, Northern America, Africa, Asia(1–6) and Australia.(7, 8) In Australia, we found that college curricula for research focus on individuals conducting their own projects and generating publications, rather than research skills development and expert supervision.(8) This current approach may encourage rushed, poor quality, small-scale projects, and trainees may fail to learn how high quality research can contribute to patient care.(9) A review of ten surgical programs in the UK highlighted similar concerns and the authors questioned the quality of the research outputs and trainee experiences.(10)

While this long-established approach to developing research skills has been questioned,(11–13) little investigation has been done to characterise the corresponding trainee research experiences and outputs. To address this information gap, we surveyed medical specialty trainees in Australia and Aotearoa/New Zealand about research activities performed as part of college training requirements. Specifically, we aimed to understand how often trainees are required to conduct research projects, how they conducted such studies, and their general views on the value of these activities. We also assessed the quality of design and reporting of their submitted research reports.

## Methods

Between April and December 2021, we conducted an anonymous cross-sectional survey of current and past medical specialty trainees. We use the CHERRIES reporting guidelines for e-surveys to report our findings.(14) Recruitment materials, full survey, and analytic code (including packages) are available on our OFS website(https://osf.io/346xe/).

### Eligibility

Eligible participants were those completing, or who had recently completed (within the past 5 years) a specialty training program at an accredited Australian or New Zealand specialty training college. No other inclusion or exclusion criteria were used.

### Recruitment

We worked with 11 medical specialty colleges to disseminate the survey via newsletters or direct email. Additional recruitment strategies included direct email with potential participants through known contacts, invitation slides at various conferences and forums, and social media posts. Potential participants were provided with a survey link and were encouraged to share the link with eligible colleagues.

The survey was distributed via an anonymous link, meaning we were unable to track which recruitment method resulted in survey participation; and were not able to provide response rates. We therefore report the number that started the survey and those who contributed to each question.

### Survey content

The survey contained three sections: a main survey (investigator developed) and two optional sections using validated instruments. We used a secure survey platform (Qualtrics(15)), with built-in survey logic so participants only saw questions relevant to them (Supplementary File 1).

The main survey section was developed by a core group of authors and was informed by the literature on known problem areas in research (16) and how best to support trainee research in the workplace.(17) It was tested for face and content validity with the wider authorship group which included experts in medical education, clinical research, research waste and Evidence-Based Practice (EBP), and representation across a wide range of specialties, and piloted with team members who included potentially eligible participants and trainee supervisors.

Participants were asked when and where they completed their most recent specialty training, their views on the importance of conducting research during specialty training, and how many projects they had completed (if any). We defined a project as any project type work that was mandated by the college as part of specialty training, including primary research, secondary research (e.g.: systematic reviews), audits and quality improvement projects.

For each project, we asked respondents: how they formulated their question; whether they performed a literature review or developed a protocol prior to commencing their project; the skills mix on the project team; access to relevant expertise and supervision; and if an how consumers were involved in the project;(18) the publication status of the project; and whether they believed the results were useful. We also asked respondents about their overall experience, including satisfaction with the overall experience, skill development opportunities, and research engagement after training.

To gain a deeper understanding of their experiences we asked trainees to complete two additional validated questionnaires. The Postgraduate Research Experience Questionnaire (PREQ) is traditionally used to assess experiences of higher degrees by research graduates across 7 domains: supervision, intellectual climate, skills development, infrastructure, thesis examination, goals and expectations, and industry engagement.(19) We excluded the industry engagement domain as our participants were based in industry. The WReN Spider instrument(20) assessed trainees’ self-perceived end-of-training research knowledge, skills and experience, focusing on the individual’s experience rather than the broader research environment.(21)

### Quality assessment of research outputs

We were unable to source trainee outputs directly from colleges, as some colleges do not archive submitted reports and others require trainee consent to release their reports. Therefore, we asked participants to upload a copy of the manuscript that was submitted to the college to the survey, or a citation of the published work. We assessed whether there was a clear question, a study rationale, adequately consideration of the published literature, and a sample size calculation (where relevant). Depending on their study type, we appraised the quality of reporting using EQUATOR guidelines(22) and the study methods using relevant critical appraisal tools (Supplementary File 2).

### Sample size

As we did not test a hypothesis, we did not undertake a formal sample size calculation taking a more pragmatic approach.(23) Using an acceptable margin of precision of ± 10% for standard prevalence estimates, and a worst-case rate of completed and uploaded research projects of 20%,(24) we estimated a sample size of around 480 responses to yield 96 completed research projects for analysis.

### Analysis

We included responses in the analyses if participants completed at least one demographic data question and analysed the survey using simple descriptive statistics. We did not conduct any sensitivity analysis or use any methods for adjusting for potential non-representativeness of our sample. Due to small response rates of trainees from individual colleges, differences in trainee responses between colleges were not explored, and factor analysis of validated surveys was not conducted. Data analysis and visualisation were conducted using Python 3.10.9(25) and R 3.6.1(26) using Jupyter notebooks(27) on a Windows 10 64-bit operating system.

Open-ended survey responses were analysed using qualitative content analysis where core meaning is derived from the text and grouped into themes.(28) This was conducted by an experienced qualitative research assistant in Microsoft Word. Themes were discussed by a subset of the team with content and qualitative research expertise (PS, CB).

### Ethics

This study was approved by the Bond University Human Research Ethics Committee (PS00149).

## Results

Of the 426 eligible participants who commenced the survey, 371 (87%) completed at least one demographic question (Figure 1), with the median time for survey completion being 5.3 minutes.

**Figure 1:**
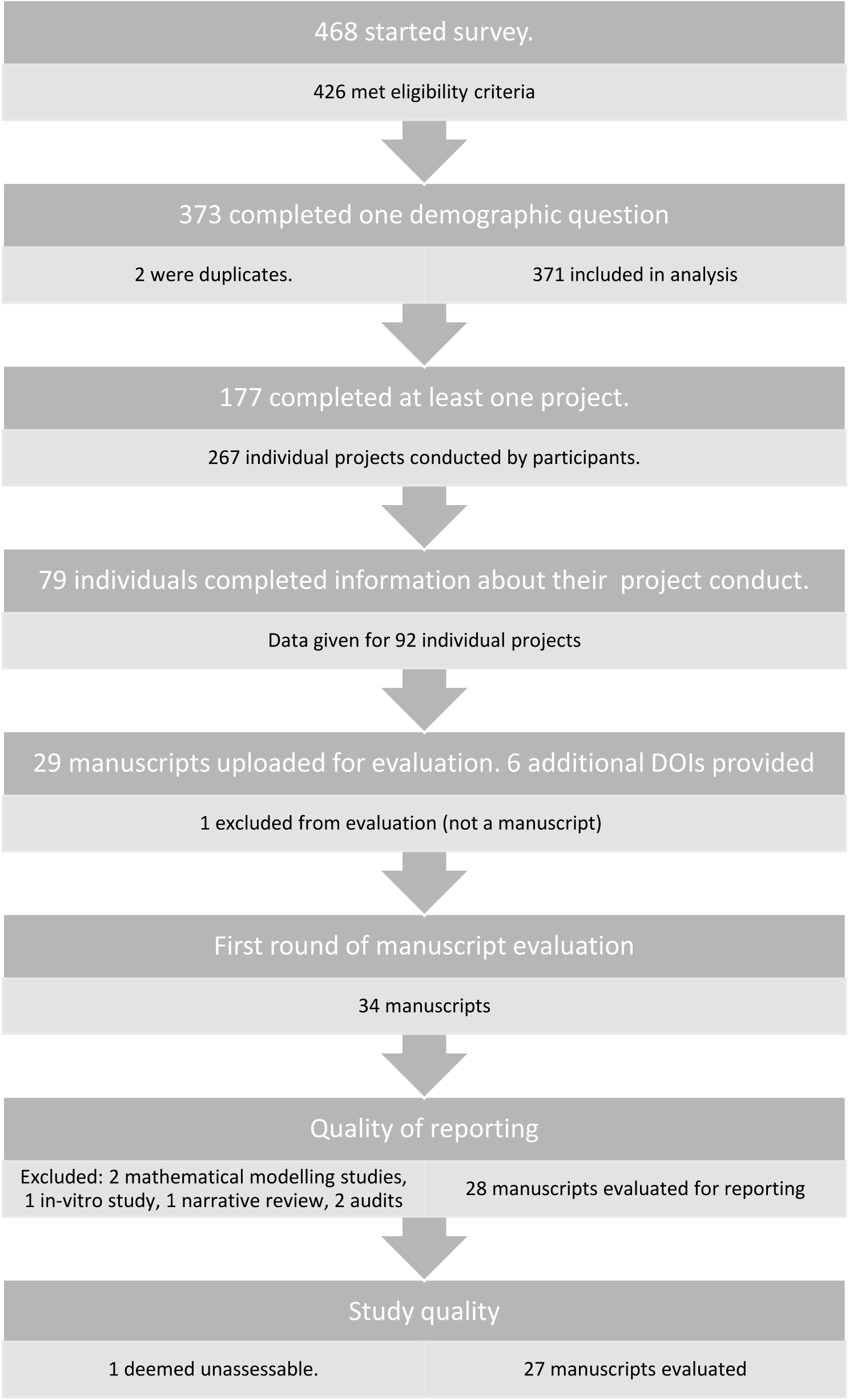
Participant data flow.

### Demographic data

Approximately two thirds of respondents were female and currently undergoing training. Of those who had completed their training, over half (58%) had finished in the previous two years. Most were undertaking their training in urban centres (Table 1). Participants represented all but one of the 16 medical specialty colleges in Australia and New Zealand (Table 1), with 323 respondents from Australia and 48 from New Zealand. Respondents from Queensland, the investigators’ home state, were over-represented (Supplementary Table 1).

**Table 1:** Demographic data. * of those eligible to answer the question. Abbreviations: RACP: Royal Australasian College of Physicians, ANZCA: Australian and New Zealand College Of Anaesthetists, RANZCP: Royal Australian and New Zealand College of Psychiatrists, RACGP: Royal Australian College of General Practitioners, ACEM: Australasian College for Emergency Medicine, CICM: College of Intensive Care Medicine, RACS: Royal Australasian College of Surgeons, RANZCOG: Royal Australian and New Zealand College of Obstetricians and Gynaecologists, RCPA: Royal College of Pathologists of Australasia, RANZCO: Royal Australian and New Zealand College of Ophthalmologists

| Answer | Number of participants | Proportion of participants |
| --- | --- | --- |
| **Gender** |  |  |
| Eligible | 371 |  |
| Total answered | 369 | 0.99* |
| Female | 224 | 0.61 |
| Male | 137 | 0.37 |
| Prefer not to say | 7 | 0.02 |
| Non-binary | 1 | 0 |
| **Specialty training college** |  |  |
| Eligible | 371 |  |
| Total answered | 371 | 1 |
| RACP | 102 | 0.27 |
| ANZCA | 86 | 0.23 |
| RANZCP | 36 | 0.1 |
| RACGP | 27 | 0.07 |
| ACEM | 26 | 0.07 |
| RACS | 23 | 0.06 |
| CICM | 22 | 0.06 |
| RANZCOG | 11 | 0.03 |
| RCPA | 9 | 0.02 |
| RANZCO | 9 | 0.02 |
| ACSEP | 5 | 0.01 |
| Other | 15 | 0.05 |
| **Completed training** |  |  |
| Eligible | 371 |  |
| Total answered | 370 | 1.0* |
| No | 237 | 0.64 |
| Post training ≤ 2 year | 77 | 0.21 |
| Post training > 2 year | 56 | 0.15 |
| **Location** |  |  |
| Eligible | 371 |  |
| Total answered | 365 | 0.98* |
| Australia | 314 | 0.86 |
| New Zealand | 48 | 0.13 |
| Other | 3 | 0.01 |
| Urban | 308 | 0.84 |
| Regional | 42 | 0.12 |
| Rural/ Remote | 15 | 0.04 |

### Research Projects

Of the 361 respondents who provided information, 311 (86%) had completed, were completing, or were planning to complete a project (Table 2). Of 47 who provided reasons for not conducting projects, 20 reported that it was not required by their college, 13 completed approved coursework and none had completed a PhD (Table 2). A third of respondents completed more than one project (48/174, 27%), equating to a total of 267 projects conducted by 173 trainees.

**Table 2:** Project completions and reasons for non-completions. * *of those eligible to answer the question*.

| Answer | Number of participants | Prop. of participants |
| --- | --- | --- |
| **Completed a project** |  |  |
| Eligible | 371 |  |
| Total answered | 361 | 0.97* |
| Yes | 177 | 0.49 |
| In progress | 76 | 0.21 |
| I plan to | 58 | 0.16 |
| No | 50 | 0.14 |
| **Reason for not completing a project** |  |  |
| Eligible | 50 |  |
| Total answered | 47 | 0.94* |
| It was not required | 20 | 0.43 |
| I had recognition of prior learning | 6 | 0.13 |
| I completed a PhD instead | 0 | 0 |
| I completed a research Masters instead | 3 | 0.06 |
| I completed approved coursework instead | 13 | 0.28 |
| Other: | 5 | 0.11 |

#### Study provenance

Of 177 trainees whose projects were completed, only 79 (45%) responded providing further information on 92 projects. Thirty-eight (41%) projects addressed questions developed by the trainees alone (Table 3). Some questions arose during clinical discussion (35/92, 38%) and a few were part of ongoing research (14/92, 15%) (Table 3). Forty-eight (52%) of study designs were developed by the trainees with little input from others. Of the 69 developed protocols, 60 were developed by the trainee and 20 were registered, including 11 in journals and 7 in registries. Most trainees searched for a systematic review of the literature prior to starting their project (68/92, 74%).

**Table 3:** Project conduct. Survey questions relevant to understanding how respondents conducted their research. * of those eligible to answer the question. †Project numbers add up to more than those that answered as each project could have answered “yes” to more than one category.

| ANSWER | NUMBER OF PROJECTS | PROP. OF PROJECTS |
| --- | --- | --- |
| **STUDY PROVENANCE** |  |  |
| **Which best describes the circumstances in which you generated your research question?** |  |  |
| Eligible | 267 | - |
| Total answered | 92 | 0.34* |
| On my own | 38 | 0.41 |
| It was a component of an ongoing project - e.g. part of a grant, one of the department priority area projects, etc | 14 | 0.15 |
| A result of a clinical discussion- e.g. recommended by my supervisor after a clinical meeting. | 35 | 0.38 |
| Other | 5 | 0.05 |
| **Which best describes the circumstances in which you generated your study design?** |  |  |
| Eligible | 267 | - |
| Total answered | 92 | 0.34* |
| On my own without or with minimal input from others. | 48 | 0.52 |
| On my own but with significant input from others. | 35 | 0.38 |
| The protocol was part of an existing project | 9 | 0.1 |
| **Before starting data collection for this project, did you develop research protocol?** |  |  |
| Eligible | 267 | - |
| Total answered | 92 | 0.34* |
| Yes - I developed one myself | 60 | 0.65 |
| Yes - there one already developed | 9 | 0.1 |
| No | 23 | 0.25 |
| **Was the protocol registered in a publicly available place? †** |  |  |
| Eligible | 69 | - |
| Total answered | 60 | 0.87* |
| Yes - Published in a journal | 11 | 0.18 |
| Yes - in a registry (e.g. PROSPERO, ClinicalTrials.gov, OSF, etc): | 7 | 0.12 |
| Yes - Other. Please state where: | 3 | 0.05 |
| No | 40 | 0.67 |
| **Before starting your project, did you search for a systematic review or other type of review (e.g. scoping review) that answered your question prior to starting your research?** |  |  |
| Eligible | 267 | - |
| Total answered | 92 | 0.34* |
| Yes | 68 | 0.74 |
| No | 24 | 0.26 |
| **RESEARCH SUPPORT REPORTED BY TRAINEES.** |  |  |

| **Did your research team consist of members outside of your own profession? †** |  |  |
| --- | --- | --- |
| Eligible | 267 | - |
| Total answered | 90 | 0.34* |
| Yes - Medical professional(s) from a different specialty.<br>Please specify: | 8 | 0.09 |
| Yes - Allied Health Professional(s). Please specify: | 10 | 0.11 |
| Yes - Nursing staff. | 10 | 0.11 |
| Yes - Statistician(s). | 21 | 0.23 |
| Yes - Health economist(s). | 1 | 0.01 |
| Yes - Librarian(s)/ Information Specialist(s). | 8 | 0.09 |
| Yes - Data scientist(s). | 5 | 0.06 |
| Yes - Other. Please specify: | 9 | 0.1 |
| No | 39 | 0.43 |
| **Please indicate if you had adequate access to any of the following types of individuals while completing your scholarly project. †** |  |  |
| Eligible | 267 | - |
| Total answered | 85 | 0.32* |
| Statistician(s) | 7 | 0.08 |
| Health economist(s) | 0 | 0 |
| Librarian(s) | 22 | 0.26 |
| Consumer or patient advocate(s) | 2 | 0.02 |
| Experts in research design or measurement | 17 | 0.20 |
| Experts in practice change strategies or practice improvement | 8 | 0.09 |
| Individuals with sufficient breadth and depth of clinical expertise | 45 | 0.53 |
| None of the above | 17 | 0.20 |
| **Your supervisor provided you with adequate research support while conducting your scholarly project.** |  |  |
| Eligible | 267 | - |
| Total answered | 85 | 0.32* |
| Strongly agree | 27 | 0.32 |
| Agree | 30 | 0.35 |
| Neither agree nor disagree | 15 | 0.18 |
| Disagree | 5 | 0.06 |
| Strongly disagree | 4 | 0.05 |
| I did not have a supervisor | 4 | 0.05 |

#### Project support and collaboration

Trainees reported low levels of interdisciplinary and interprofessional collaboration, with 43% (39/90) of project teams consisting of only members of their own specialty. Forty-four percent (40/90) of project teams consisted of only one other profession, often a statistician, allied health professional, or nurse, but seldom other disciplines (Table 3). Most respondents (68/85, 80%) reported obtaining some expert support – most commonly clinical expertise, library services and study design or measurement expertise (Table 3). Only 7 out of 90 (8%) projects involved consumers (Supplementary Table 2). Most (57/85, 67%) reported that they received adequate support from their supervisor. Trainees from all but two colleges reported carrying out the research work in their own time (Table 4). The PREQ survey explored research support further. Only 10 responded (Supplementary Figure 1). Respondents were least satisfied with the intellectual climate in which they conducted research and opportunities for skills development, and most satisfied with the thesis examination process and understanding project requirement goals and expectations.

**Table 4:** When projects were conducted. We asked participants to estimate the percentage of time they spent on their scholarly projects during scheduled service/clinical time, protected time or during their own time. We received responses for 85/267 projects. Red highlights indicate highest median number for that row. RACGP’s trainee program offers an academic post which provides funding for protected research time. * 5 participants gave information on 10 projects, † 1 participants gave information on 1 project, ‡2 participants gave information on 4 projects, §1 participant gave information on 7 projects. Abbreviations: ACEM: Australasian College for Emergency Medicine, ACSEP: Australasian College of Sport and Exercise Physicians, ANZCA: Australian and New Zealand College Of Anaesthetists, CICM: College of Intensive Care Medicine, RACDS: Royal Australasian College of Dental Surgeons, RACGP: Royal Australian College of General Practitioners, RACMA: Royal Australasian College of Medical Administrators, RACP: Royal Australasian College of Physicians, RACS: Royal Australasian College of Surgeons, RANZCOG: Royal Australian and New Zealand College of Obstetricians and Gynaecologists, RANZCP: Royal Australian and New Zealand College of Psychiatrists, RANZCR: Royal Australian and New Zealand College of Radiologists, RCPA: Royal College of Pathologists of Australasia

| College | Number of projects | Clinical time (%) |  | Protected time (%) |  | Own time (%) |  |
| --- | --- | --- | --- | --- | --- | --- | --- |
|  |  | Median | IQR | Median | IQR | Median | IQR |
| ACEM | 2 | 5 | 2 - 7 | 5 | 2 - 7 | 90 | 90 - 90 |
| ACSEP | 2 | 28.5 | 24 - 32 | 15.5 | 7 - 23 | 56 | 44 - 68 |
| ANZCA | 20* | 7 | 3 - 16 | 0 | 0 - 1 | 90 | 71 - 95 |
| CICM | 10 | 4.5 | 0 - 5 | 0 | 0 - 3 | 92.5 | 89 - 98 |
| RACDS | 2† | 25 | 25 - 25 | 0 | 0 - 0 | 75 | 75 - 75 |
| RACGP | 3 | 5 | 2 - 6 | 85 | 80 - 89 | 10 | 8 - 14 |
| RACP | 14‡ | 8 | 0 - 10 | 6 | 0 - 20 | 81 | 71 - 94 |
| RACS | 8 | 5 | 4 - 16 | 2.5 | 0 - 27 | 86 | 53 - 95 |
| RANZCOG | 1 | 0 | 0 - 0 | 0 | 0 - 0 | 100 | 100 - 100 |
| RANZCP | 13 ‡ | 10 | 0 - 11 | 19 | 0 - 25 | 75 | 52 - 89 |
| RANZCR | 3 | 0 | 0 - 1 | 0 | 0 - 1 | 100 | 98 - 100 |
| RCPA | 7§ | 90 | 7 - 100 | 0 | 0 - 0 | 10 | 0 - 92 |
| Overall | 85 | 5 | 0 - 16 | 0 | 0 - 11 | 89 | 66 - 95 |

#### Perceived value of the research findings and dissemination of results

Most participants (78/92, 88%) indicated the results of their study would be useful in practice (78/92, 88%), and the majority (81/90, 90%) felt confident using the results in practice. (Table 5). Half of the projects (46/90, 51%) have a publicly available manuscript. Of those published in a journal, the trainee was usually first author (37/42, 88%) and approximately a third (33/90, 37%) were published by the end of training.

**Table 5:** Project value. * of those eligible to answer the question.

| ANSWER | NUMBER OF PROJECTS | PROP. OF PROJECTS |
| --- | --- | --- |
| **PERCEIVED VALUE OF RESEARCH FINDINGS AND PUBLICATION STATUS** |  |  |
| **Do you or your colleagues believe that the results of this study may be useful in practice?** |  |  |
| Eligible | 267 | - |
| Total answered | 89 | 0.33* |
| Yes | 78 | 0.88 |
| No | 11 | 0.12 |
| **How confident are you in using the findings of your study in clinical practice?** |  |  |
| Eligible | 267 | - |
| Total answered | 90 | 0.34* |
| Very confident | 39 | 0.43 |
| Somewhat confident | 42 | 0.47 |
| Not at all confident | 9 | 0.1 |
| **Is a manuscript containing the results publicly available?** |  |  |
| Eligible | 267 | - |
| Total answered | 90 | 0.34* |
| Yes - Published in a journal by the end of your training | 33 | 0.37 |
| Yes - Subsequently published in a journal | 12 | 0.13 |
| Yes - Pre-print available | 1 | 0.01 |
| No - It is unpublished | 44 | 0.49 |
| **Which author position did you have for this publication?** |  |  |
| Eligible | 45 | - |
| Total answered | 42 | 0.93* |
| First | 37 | 0.88 |
| Second | 4 | 0.1 |
| Last | 1 | 0.02 |
| Other | 0 | 0 |
| **PERCEIVED PERSONAL VALUE OF CONDUCTING RESEARCH PROJECTS** |  |  |
| **How important did you feel conducting a scholarly project was to your clinical career development?** |  |  |
| Eligible | 177 |  |
| Total answered | 174 | 0.98* |
| Very important | 30 | 0.17 |
| Moderately important | 57 | 0.33 |
| Slightly important | 56 | 0.32 |
| Not at all important | 31 | 0.18 |
| **I had the necessary knowledge and skills to complete my scholarly project(s).** |  |  |
| Eligible | 177 |  |
| Total answered | 68 | 0.38* |
| Strongly agree | 12 | 0.18 |
| Agree | 26 | 0.38 |
| Neither agree nor disagree | 15 | 0.22 |
| Disagree | 14 | 0.21 |
| Strongly disagree | 1 | 0.01 |
| **Completing a scholarly project(s) during my specialty training gave me a better understanding of how to read and interpret other people's research** |  |  |
| Eligible | 177 |  |
| Total answered | 68 | 0.38* |
| Strongly agree | 17 | 0.25 |
| Agree | 23 | 0.34 |
| Neither agree nor disagree | 9 | 0.13 |
| Disagree | 13 | 0.19 |
| Strongly disagree | 6 | 0.09 |
| **Since gaining your most recent fellowship, have you considered initiating a new research project?** |  |  |
| Eligible | 133 |  |
| Total answered | 61 | 0.46* |
| Yes | 44 | 0.72 |
| No | 17 | 0.28 |
| **Since gaining your most recent fellowship, have you participated in any research projects as an investigator?** |  |  |
| Eligible | 133 |  |
| Total answered | 61 | 0.46* |
| Yes | 33 | 0.54 |
| No | 28 | 0.46 |

#### Respondents’ views on mandatory research projects

Almost half the participants who completed a project (87/174, 50%) felt that this effort was very or moderately important to their clinical career and over half (40/68, 59%) felt that completing a research project improved their ability to read and interpret research (Table 5).

The participants responses on the value of conducting mandatory research projects were mixed, with around half of the respondents expressing positive attitudes (Supplementary Table 3). When asked why, 236 participants provided a response. Sixty-five (27%) participants mentioned the time required to do the research was unreasonable given clinical workloads and time away from family life and other priorities (Table 6). Fifty-one participants (21%) felt mandatory projects contributed to poor quality research and 21 (9%) described them as “tick box” activities. Thirty-nine (17%) participants described a lack of structured support in the current training program, 36 (15%) suggested the research projects were a waste of time or not relevant to their career objectives, and 28 (12%) suggested there were better ways to learn EBP or research skills. While 29 (12%) recommended research should be optional rather than mandated, 44 (19%) participants suggested mandatory projects were important to develop skills beyond just research, 18 (8%) suggested it improved their EBP skills and 14 (6%) suggested they improved clinical practice.

**Table 6:** Codes from free-text content analysis.

| Theme | ILLUSTRATIVE QUOTES |
| --- | --- |
| Why trainees they supported/opposed mandatory projects. |  |
| Important to skills development including EBM. Improves practice. Is important.<br>(n = 76) | *"Completion of this task is a good test of organisation, prioritisation irrespective of the research component. All medical staff should be able to critical[ly] appraise literature." (Strongly supports mandatory projects)*<br>*"I am not personally strongly interested in pursuing research as part of my career but recognise that it is unavoidable in modern medicine and required as part of any job application." (Moderately supports mandatory projects)*<br>*"Think it has some value for learning and rounding of a physician's skills." (Moderately supports mandatory projects)* |
| Contribution to research waste. Tick box activity.<br>(n = 72) | *"To require true research for entrance or progression does nothing more than produce rubbish research." (Neither supports nor opposes mandatory projects)*<br>*"I think in general mandatory research requirement to produce "papers" contribute to a large bubble of generally irrelevant papers which adds to a constant background of research noise that doesn't actually change practice." (Moderately opposes mandatory projects)*<br>*"There is already an abundance of very questionable registrar level "research" diluting the pool of genuine, high quality, and clinically useful publications that are available. Cynically completing a research project because you are force[d] to do so does not benefit the individual or the profession, rather the opposite. (Strongly opposes mandatory projects)* |
| Time. Unreasonable time requirements. Time away from life. Other prioritises.<br>(n=65) | *"While knowing how to publish is a good thing for trainees the practical experience of it I prohibitive and can delay completion of your training." (Moderately supports mandatory projects)*<br>*"Whilst I strongly support research in general and feel the experience is beneficial personally I feel the requirement to carry out compulsory, time-consuming research unpaid and with no allocated time whilst working more than full time and completing other training requirements and attending to family etc is unethical and needs to be reconsidered by all colleges" (Moderately opposes mandatory projects)* |
| No structural support.<br>(n = 39) | *"Almost the entire project is done in my spare time, this ended up being hundreds of hours... there is no access to any kind of research resources by the college, other than a handful of PDFs of previous* |
|  | *projects on the website. It's a great idea, but as a trainee, I am tired of being forced to spend my spare time outside of work (when I should be relaxing/having a family/doing hobbies) devoted to mandatory training that is not supported by the college. We are stuck doing boring projects... on our own time, and end up with the worst of both worlds."* (Strongly supports mandatory projects)<br>*"I do think research experience is important, but more support and guidance should be provided by colleges to meet their expectations. I had a disinterested supervisor (who I had to find myself) and a statistician who went on holiday for 5 months without telling me! It was a nightmare."* (Moderately supports mandatory projects)<br>*"Without formalised and adequate oversight by knowledgeable staff, the quality of such endeavors is often poor... both projects were completed with essentially no outside input/help, so I can't really speak for the statistical quality or relevance of either."* (Neither supports nor opposes mandatory projects)<br>*"Not enough support, guidance or time provided for project work. It is extremely difficult to find time to complete your project as well as an appropriate supervisor with the time, interest and experience in research."* (Strongly opposes mandatory projects) |
| Not relevant – waste of time.<br>(n = 36) | *"Most of us won't end up in research roles so while I feel that it's imperative that we know how to effectively interpret evidence, I don't think it's a great use of time to mandate research projects for trainees that don't have a particular interest in that area."* (Neither supports nor opposes mandatory projects)<br>*"Seems a waste of time on the whole. Most of the "research" done as compulsory research for training isn't proper research, contributes little if anything to the field and doesn't teach the person doing it anything about real research (I say this having done proper research prior to medicine)"* (Moderately opposes mandatory projects)<br>*"It is unnecessary for clinical work. Medical research should be its own specialty. There is so much crap research that is performed for the sake of it. It is a waste of time."* (Strongly opposes mandatory projects) |
| Optional, not mandated<br>(n = 29) | *"I think completing a scholarly project teaches essential skills in evidence-based medicine and critical appraisal. However, I also acknowledge that not all doctors are interested in research and I don't think it should be made compulsory."* (Moderately support mandatory projects) |
|  | *"I can see some virtue to this, but the implication that every specialist has to be a researcher is invalid. Additionally, the requirement to conduct your own study and be first author (as opposed to participating in a multi-centre study) excludes a lot of good experience and encourages poor practices." (Neither supports nor opposes mandatory projects)* |
| Better ways to learn these skills (EBM and otherwise)<br>(n = 28) | *"There are other ways of developing research skills, particularly for those who have little or no interest in an academic path. For example, journal clubs, unit/departmental meetings." (Neither supports nor opposes mandatory projects)*<br>*"It is valuable to participate in research though and to learn the finer points and have better understanding of the process. It would perhaps be more valuable to assess the quality of the project participated in and the contribution rather than the first author status." (Moderately opposes mandatory projects)*<br>*"The skill in interpreting research is much better taught in an academic environment rather than forcing people without any background in research to complete often low-quality research in an unsupported manner. (Strongly opposes mandatory projects)* |
| Challenges with project (moving hospital, bureaucracy) (n = 9) | *"I think it is very hard to complete the research project when the requirements of training mean short contracts and constantly moving hospitals and states. Without a longer-term engagement with one centre it is hard to be involved in meaningful research. (Moderately opposes mandatory projects)* |
| **Reasons trainees conducted research after their training:** |  |
| Have time or interest in research. Believe that research is important. Opportunities provided.<br>(n = 41) | *"I am in a role which allows non-clinical time to achieve these goals."*<br>*"Because I am still passionate about [my research area] and it is an important part of my job."* |
| Another training program. (n = 9) | *"Had to do [a project] during my own subspecialty fellowship"* |
| Supporting others/trainees (n = 5) | *"I'm involved in other trainees' projects due to my skills."* |
| **Reasons trainees did not conduct research after their training:** |  |
| Other prioritises, no time. Not interested. (n = 24) | *"I enjoy research, but again, very difficult to fit in whilst working full time. This is particularly true as a consultant."* |
| No opportunities, supports. Interested, but... (n= 13) | *"Lack of funding or pathways to continue research"* |

Self-perceived end-of-training research skills were explored further using the WReN Spider instrument. Only 10 responded (Supplementary Figure 2). All respondents felt they were somewhat to very experienced in finding and appraising the literature, and less than half felt this way about protocol writing, publishing, qualitative methods and acquiring funding.

Since completing their training, almost two thirds of respondents (44/61, 72%) had thought about initiating new research after completing their training, and approximately half had participated in research (Table 5). When asked the reason for their answers in free text, 36/56 (64%) participants commented they now had more time and interest to participate in research, whereas 21/56 (38%) said they had no time (Table 6).

#### Research outputs – quality of methods and reporting

Respondents uploaded 34 studies (Supplementary Table 4); 28 were assessed for quality as six were excluded due to a lack of standardised instruments.

Overall, the introduction and discussion sections were well reported; however, there were gaps in reporting in other sections (Figure 2). Most studies had moderate to high risk of bias; 3/27 were deemed to have low risk of bias and one was unassessable (Supplementary Figure 3).

**Figure 2:**
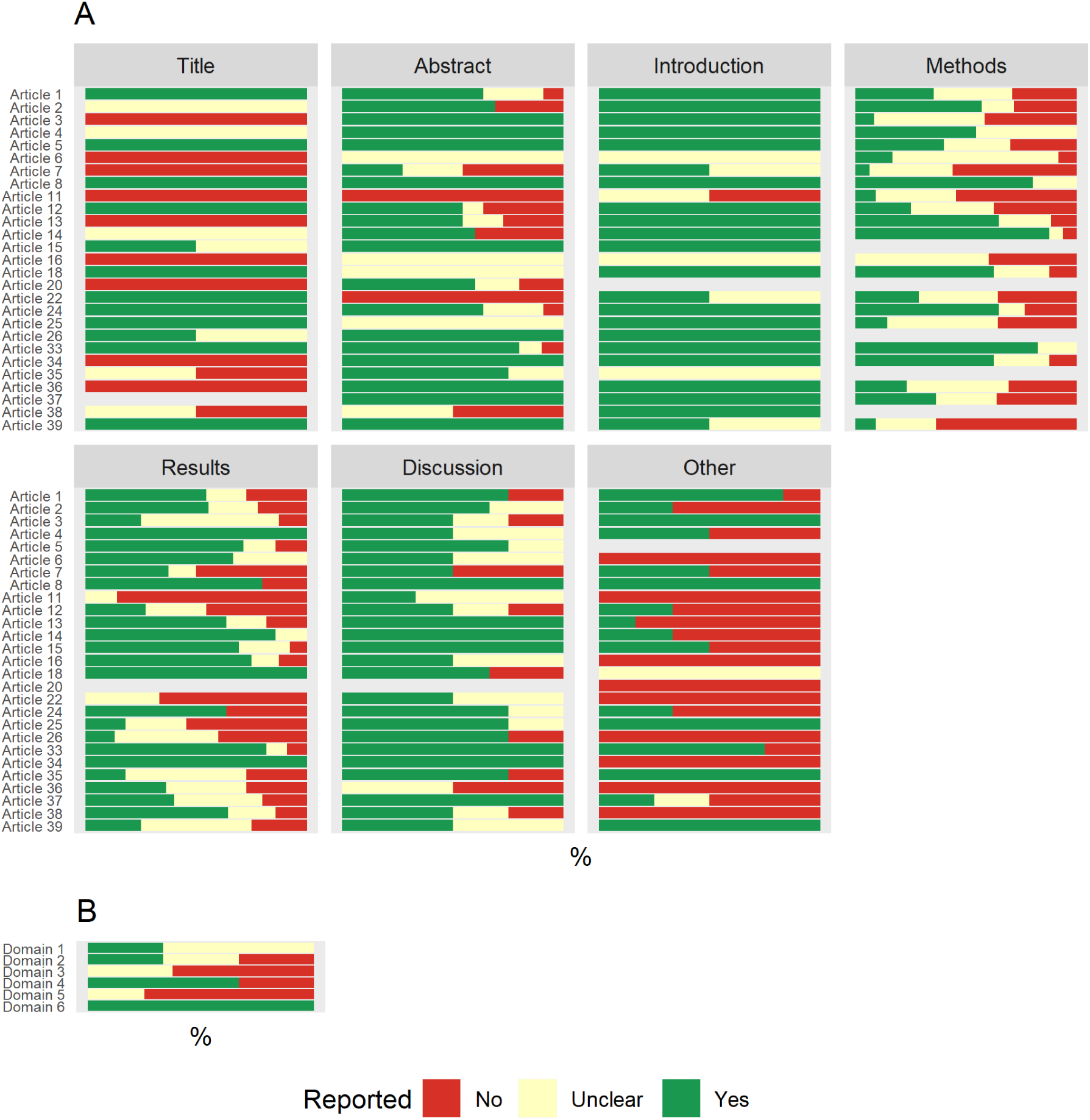
Quality of Reporting. Panel A contains 27/28 evaluated manuscripts. Each line represents an uploaded manuscript. One manuscript was a clinical guideline and did not map to same sections (i.e.: title, abstract, introduction, methods, results, discussion and other) and can be found in Panel B. Some lines are blank because they were an abstract only (Article 20) or because CARE reporting guidelines do not have a methods section (Articles 15, 26, 35, 38) and the ARRIVE reporting guideline (Article 37) merges title and abstract together.

## Discussion

Most trainees in this study were required to complete a research project as part of the specialty training. Overall, responses indicated that educational experiences and quality of research outputs were inconsistent. However, in our view the most significant finding of our study was the significant attrition of responses at each stage of the survey. Of 177 trainees who reported completion of a research project, just over one third of trainees responded to questions regarding how they conducted their project, and one sixth uploaded a project. We feel that those who had a better experience were more likely to respond providing a more positive picture than might be observed more broadly. Despite this, our results are enlightening.

Approximately half of the respondents were solely responsible for developing their research questions, designing their studies, and developing study protocols, while only few projects were part of ongoing research. Although most reported adequate support from their project supervisors, they worked in non-collaborative teams often with only their own specialty. Although statisticians, allied health or nurses were frequently represented in research teams, trainees reported low levels of access to additional expertise. This may reflect lack of research opportunities and resources or the view that medical specialists are expected to learn by doing and leading research irrespective of their baseline abilities.

Most conducted projects in their own time. Exceptions to this were those from the Royal Australian College of General Practitioners, which provide protected time for 20 trainees per year and don’t require a project from others, and the Royal College of Pathologists of Australia, which conducted most of their research during clinical time.

When we assessed uploaded projects’ design and reporting quality, reporting standards were met to a fair degree, and few reports had study methods judged to have a low risk of bias.

The trainees who provided details of their completed projects reported some positive features. Most searched the literature for systematic reviews before starting, two thirds drafted research protocols and over one third were registered. Half of the project reports were published in journals, usually prior to respondents completing their training. Most thought their project’s findings were useful and likely to use them in clinical practice, and that the experience of conducting the research project was important to their career.

Negative views were expressed by those both supportive and unsupportive of mandatory projects including conducting projects uncompensated in their own time, competing with family commitments; a lack of structured support; and concerns their projects were ‘tick box’ projects which simply contributed to unhelpful research findings - in other words, research waste. Respondents also commented that learning how to apply research evidence in practice as preferable to conducting projects.

Our study has limitations. While we are unable to judge the true representativeness of this sample, our results are likely biased towards more positive experiences. The uploaded project reports had a higher publication rate compared to the broader study cohort and in health and medical literature more generally (74% v 50%) (29–31), likely representing better quality studies than the broader trainee population. Since completing their training, over 70% of respondents had considered participating in research and over half had become involved in research, figures much higher than previously reported for Australian specialist medical practitioners.(32) This suggests our respondents may have a higher interest in research than the broader trainee population. There is probably a large silent majority who withheld generally negative views.

The real value of this educational approach can be judged by asking the right questions: ‘What do we aim to achieve by providing research training to doctors? And what is the best way to get there?’ While government reports suggest the need for better translation of research into practice and familiarity with contemporary research methods,(33) clear strategies for achieving these goals have not been clearly enunciated by the relevant professional bodies.

Most will agree that every practitioner should be competent in translating research findings into their practice; however, requiring every trainee to undertake a research project to teach EBP is not fit for purpose.(34) Some, like the Royal Australian College of General Practitioners and Australasian College of Emergency Medicine, have recognised this.(12, 35) At present less than 1% of Australian doctors identify as being a researcher, and less than 8% participate in research.(32, 36). The small number of trainees who go on to be research leaders will be internally driven to do so and should be well supported from an early stage. This leaves a substantial number who could contribute to worthwhile collaborative research enterprises (e.g., participation in large adaptive platform trials and observational studies) but who are not currently being prepared for this activity.

Trainee research collaboratives (TRC) are a potential avenue to learn such skills. These have been used in the United Kingdom since 2007 and produce high quality research while providing developing trainees skills. (37, 38) TRCs are beginning to form across Australia and New Zealand,(39) but unless trainees are first authors, contributions rarely receive college recognition.

We believe there are two important unintended consequences of this well-meaning tradition of leading research for specialist qualification. First is the likely contribution to the wider issue of research waste though poorly planned and executed projects. However, being able to support every trainee to lead a study that meaningfully contributes to the scientific body of literature takes substantial resourcing that is neither feasible nor sustainable. The second, and perhaps more significant implication, is the missed opportunity when trainees are tasked with leading research instead of honing research skills relevant to their career objective – which, for most trainees is to be an evidence-based clinicians - and to prepare clinicians for collaborative research. Future work should articulate a minimum set of research competencies and develop a flexible training curriculum that can be adapted to the career needs and aspirations of individuals.

## Data Availability

All data produced in the present study are available upon reasonable request to the authors.

https://osf.io/346xe/

## Contributions, guarantor information and acknowledgements

All authors agree with the viewpoints expressed in this manuscript. PS DH CB and CN conceptualised the study. PS, CW, RB, AB, CB, CN, PG, IS, ABB, MM, GK, HJ, EV, KF, DP, SM, DH as well as David Ellwood, David Pearson, Rhea Liang, Gordon Wright all contributed to the development of the methods and funding acquisition. PS developed the formal analysis plan (with guidance from AB), developed the data extraction tools, data curation processes, validation processes, conducted the data wrangling and formal analysis, developed the analytic and visualisation code, and oversaw project administration. She is the study guarantor. All listed authors contributed to dissemination of the survey, which was managed by Iris Gerke and overseen by PS. All authors contributed to the data extraction of uploaded files. We would like to acknowledge Tammy Hoffman and Mina Bakhit for their advice on using reporting guidelines, and Joanne Hilder for conducting the content analysis. PS developed the original draft of this manuscript, DH CB and AB provided initial critical reviews, and all authors reviewed and approved the final version of this manuscript.

The corresponding author attests that all listed authors meet authorship criteria and that no others meeting the criteria have been omitted.

## Supplementary Files

### Supplementary File 1: Additional information on survey procedure

Participants could access the survey via the Qualtrics platform. Incomplete responses were recorded. Respondents were also prevented from multiple submissions (settings used on Qualtrics system: “This setting works by placing a cookie on their browser when they submit a response.”)

Section 1 (all questions mandatory except project file upload) of the survey was around 10 pages long (depending on responses). The PREQ and WReN Spider instrument were one page each. Based on piloting feedback and a desire to reduce the burden on trainees, these sections were not mandatory, and participants were asked if they wished to provide data on these before being shown these sections.

Participants were able to use the “back” button to change their responses and were able to come back to surveys and finish them off up to 3 months after closing their browser before being recorded.

At the beginning of the survey, participants provided some information to create a unique identifier based their initials, day and month of birth, and gender. This was used to check for duplicates and remove surveys. Zero participants requested that their data be removed. Potential duplicate responses were examined manually, and two were identified as duplicates, their second response was included for analysis.

Participation was completely voluntary, and no incentives were offered to participants.

### Supplementary File 2: Quality assessment of research outputs

We assessed each uploaded project through two rounds of data extraction. During the first round we categorised the submission type, research question type, study design, and whether the upload was an audit. We assessed whether the authors asked a clear question, provided a study rationale, adequately considered the published literature, or provided a sample size calculation (where relevant). During data extraction we noticed that many projects did not explicitly label, or mislabelled, the study design; we therefore added this as an extra variable during data extraction. For published manuscripts we checked whether the journal stated they used a peer-review process, and whether the journal was listed on the Predatory Journal list.(1)

During the second round of data extraction, we assessed the quality of reporting and design of each upload. We used EQUATOR-network(2) reporting guidelines to assess the quality of reporting of individual studies (*below*). We had originally planned to use risk of bias tools recommended by Cochrane to assess study quality, however, due to the wide variety of study questions and designs, we felt it would be difficult to interpret results from several different tools and we therefore modified them as described in the Table below.

Given that some uploads mislabelled their study design or did not provide a study design we used the following rules to decide on the quality assessment tool. For studies that incorrectly labelled their study, we assessed the quality of reporting based on the study design they assigned themselves and the design quality assessment on the actual study design used. For those that did not provide a study design label we assigned the study design based on information given in their methods section and used the relevant reporting and design quality tools. We excluded studies from quality assessment where a reporting guideline or critical appraisal instrument was not available.

All data extraction was done independently by two authors. Discrepancies were resolved by discussion, or by discussion with a third author with the relevant skill set (AB was used for all statistical resolutions and DH and PG were used for all methodological/design resolutions).

**Table:** Reporting and design quality assessment tools. * We had 1 animal study which was a randomised trial and could therefore be evaluated using the MMAT quantitative randomized controlled trial tool. †The AMSTAR signalling questions were modified slightly to allow for evaluation of non-interventional studies and scoping reviews. This was done with 3 team members, 1 of which is an author on the original AMSTAR tool (DH), a statistician (AB), and systematic review expert (ABB). ‡ Risk of bias elements only.

| TOOL USED | STUDY DESIGNS |
| --- | --- |
| **Reporting Quality** |  |
| AGREE(3) | Guidelines |
| ARRIVE(4) | Animal studies |
| CARE(5) | Case reports, Case Series |
| CONSORT Cross-over(6) | Cross over randomised trial |
| PRISMA (7) | Systematic Reviews |
| PRISMA-ScR (8) | Scoping Reviews |
| SRQR (9) | Qualitative |
| STARD (10) | Diagnostic test accuracy |
| STROBE (11) | Cohort, Case control, Cross Sectional |
| **Design Quality** |  |
| MMAT(12) | Animal studies*, Case reports, Case Series, Cross over randomised trial, Qualitative, Cohort, Case control, Cross Sectional |
| Modified AMSTAR-2 (13)† | Systematic Review, Scoping Review |
| QUADAS-2 (14)‡ | Diagnostic test accuracy |
| MiChe (15) | Guidelines |

### Supplementary Tables

**Supplementary Table 1:** Additional demographic data. * of those eligible to answer the question. † appears as zero due to rounding. Abbreviations: ACD: Australasian College of Dermatologists, ACEM: Australasian College for Emergency Medicine, ACRRM: Australian College of Rural and Remote Medicine, ACSEP: Australasian College of Sport and Exercise Physicians, ANZCA: Australian and New Zealand College Of Anaesthetists, CICM: College of Intensive Care Medicine, RACDS: Royal Australasian College of Dental Surgeons, RACGP: Royal Australian College of General Practitioners, RACMA: Royal Australasian College of Medical Administrators, RACP: Royal Australasian College of Physicians, RACS: Royal Australasian College of Surgeons, RANZCO: Royal Australian and New Zealand College of Ophthalmologists, RANZCOG: Royal Australian and New Zealand College of Obstetricians and Gynaecologists, RANZCP: Royal Australian and New Zealand College of Psychiatrists, RANZCR: Royal Australian and New Zealand College of Radiologists, RCPA: Royal College of Pathologists of Australasia

| ANSWER | NUMBER OF PARTICIPANTS | PROP. OF PARTICIPANTS |
| --- | --- | --- |
| **Specialty training college** |  |  |
| Eligible | 371 |  |
| Total answered | 371 | 1 |
| ACD | 3 | 0.01 |
| ACEM | 26 | 0.07 |
| ACRRM | 2 | 0.01 |
| ACSEP | 5 | 0.01 |
| ANZCA | 86 | 0.23 |
| CICM | 22 | 0.06 |
| RACDS | 1 | 0† |
| RACGP | 27 | 0.07 |
| RACMA | 0 | 0 |
| RACP | 102 | 0.27 |
| RACS | 23 | 0.06 |
| RANZCO | 9 | 0.02 |
| RANZCOG | 11 | 0.03 |
| RANZCP | 36 | 0.1 |
| RANZCR | 5 | 0.01 |
| RCPA | 9 | 0.02 |
| OTHER | 2 | 0.01 |
| Prefer not to say | 2 | 0.01 |
| **Year training was completed** |  |  |
| Eligible | 133 |  |
| Total answered | 133 | 1.0* |
| 2015 | 2 | 0.02 |
| 2016 | 11 | 0.08 |
| 2017 | 22 | 0.17 |
| 2018 | 21 | 0.16 |
| 2019 | 22 | 0.17 |
| 2020 | 55 | 0.41 |
| **Which country and state did you complete/ are completing most of your most recent specialty training in?** |  |  |
| Eligible | 371 |  |
| Total answered | 365 | 0.98* |
| Australia: Australian Capital Territory | 3 | 0.01 |
| Australia: New South Wales | 75 | 0.2 |
| Australia: Northern Territory | 2 | 0.01 |
| Australia: Queensland | 102 | 0.27 |
| Australia: South Australia | 16 | 0.04 |
| Australia: Tasmania | 4 | 0.01 |
| Australia: Victoria | 74 | 0.2 |
| Australia: Western Australia | 38 | 0.1 |
| New Zealand: - | 48 | 0.13 |
| Other: - | 3 | 0.01 |

**Supplementary Table 2:** Additional information on study conduct. * *Number of participants. † of those eligible to answer the question*.

| ANSWER | NUMBER OF PROJECTS | PROP. OF PROJECTS |
| --- | --- | --- |
| **COLLEGES REPRESENTED IN THE DATA** |  |  |
| **Colleges that gave project data** |  |  |
| Eligible | 177* |  |
| Total answered | 79* | 0.45† |
| ACD | 0 | 0 |
| ACEM | 2 | 0.03 |
| ACRRM | 0 | 0 |
| ACSEP | 2 | 0.03 |
| ANZCA | 18 | 0.23 |
| CICM | 11 | 0.14 |
| RACDS | 1 | 0.01 |
| RACGP | 3 | 0.04 |
| RACMA | 0 | 0 |
| RACP | 16 | 0.2 |
| RACS | 8 | 0.1 |
| RANZCO | 0 | 0 |
| RANZCOG | 1 | 0.01 |
| RANZCP | 11 | 0.14 |
| RANZCR | 2 | 0.03 |
| RCPA | 4 | 0.05 |
| OTHER | 0 | 0 |
| **STUDY PROVENANCE** |  |  |
| **Were consumers involved in the design of your research?** |  |  |
| Eligible | 267 | - |
| Total answered | 90 | 0.34† |
| Yes | 7 | 0.08 |
| No | 83 | 0.92 |
| **Which part of the research process the consumers were involved in?** |  |  |
| Eligible | 7 | - |
| Total answered | 7† | 1.0† |
| Developing the research question | 0 | 0 |
| Protocol design | 3 | 0.43 |
| Conduct of research | 6 | 0.86 |
| Dissemination of research | 2 | 0.29 |
| Future work including implementation of research findings and/or developing future research questions | 3 | 0.43 |
| **Which part of the research process the consumers were involved in?** |  |  |
| Eligible | 7 | - |
| Total answered | 7 | 1.0† |
| Consultation | 7 | 1 |
| Co-investigator/collaborator | 0 | 0 |
| Lead | 0 | 0 |
| **RESEARCH SUPPORT REPORTED BY TRAINEES.** |  |  |
| **I had access to a good research-related seminar(s) or training program.** |  |  |
| Eligible | 177 |  |
| Total answered | 68 | 0.38<sup>†</sup> |
| Strongly agree | 6 | 0.09 |
| Agree | 13 | 0.19 |
| Neither agree nor disagree | 8 | 0.12 |
| Disagree | 28 | 0.41 |
| Strongly disagree | 13 | 0.19 |

**Supplementary Table 3:** Project value - additional details.

| ANSWER | NUMBER OF PROJECTS | PROP. OF PROJECTS |
| --- | --- | --- |
| **PERCEIVED VALUE OF RESEARCH FINDINGS AND PUBLICATION STATUS** |  |  |
| **Were the results of your study presented to the department where are doing or did your clinical training?** |  |  |
| Eligible | 267 | - |
| Total answered | 90 | 0.34* |
| Yes | 68 | 0.76 |
| No | 22 | 0.24 |
| **PERCEIVED PERSONAL VALUE OF CONDUCTING RESEARCH PROJECTS** |  |  |
| **Overall, I was satisfied with the quality of my research experience during my specialty training** |  |  |
| Eligible | 177 |  |
| Total answered | 68 | 0.38* |
| Strongly agree | 13 | 0.19 |
| Agree | 17 | 0.25 |
| Neither agree nor disagree | 17 | 0.25 |
| Disagree | 15 | 0.22 |
| Strongly disagree | 6 | 0.09 |
| **How much do you support or oppose the requirement to complete a scholarly project during specialty training?** |  |  |
| Eligible | 371 |  |
| Total answered | 67 | 0.18* |
| Strongly support | 19 | 0.28 |
| Moderately support | 17 | 0.25 |
| Neither support nor oppose | 7 | 0.10 |
| Moderately oppose | 14 | 0.21 |
| Strongly oppose | 10 | 0.15 |

**Supplementary Table 4:** Uploaded project demographics. This provided an overall picture of the kinds of studies trainees were conducting as part of their specialty training research requirements. * Two individuals uploaded to articles each. †All four articles were from the same individual. ‡ One published in a potentially predatory journal. § “Ad-hoc” here meant that it was not made clear in the manuscript that the audit or quality improvement project was part of a pre-specified local or state/national project. ¶ This upload was a poster, and we did not feel providing a comprehensive overview of the literature was applicable.

| QUESTION | CATEGORY | NUMBER OF ARTICLES |
| --- | --- | --- |
| **DEMOGRAPHIC DATA** |  |  |
| **College** |  |  |
|  | RANZCP | 8 (24%) |
|  | CICM | 7 (21%) |
|  | ANZCA* | 7 (21%) |
|  | RCPA† | 4 (12%) |
|  | RACS | 3 (9%) |
|  | RACP | 2 (6%) |
|  | RACGP | 1 (3%) |
|  | RANZCOG | 1 (3%) |
|  | ACEM | 1 (3%) |
| **What type of submission was provided?** |  |  |
|  | Published Manuscript‡ | 25 (74%) |
|  | Full Text manuscript/ report | 8 (24%) |
|  | Poster | 1 (3%) |
| **Was this an Audit or QI project?** |  |  |
|  | No | 32 (94%) |
|  | Yes - AD-HOC§ | 2 (6%) |
| **WERE THE QUESTIONS RELEVANT TO USERS OF RESEARCH?** |  |  |
| **Did the authors provide a sound argument for the rationale to do the study and/or that the results of the study provide meaningful information?** |  |  |
|  | Yes | 31 (91%) |
|  | No | 3 (9%) |
| **Was there an adequate consideration of the published literature on the topic, including previous systematic reviews?** |  |  |
|  | Yes | 23 (68%) |
|  | No | 10 (29%) |
|  | N/A ¶ | 1 (3%) |
| **Was there a clear, well-structured, and answerable research question? (e.g. PICO-T).** |  |  |
|  | Yes | 28 (82%) |
|  | No | 4 (12%) |
|  | Partial | 2 (6%) |
| **What type of question did the researchers ask?** |  |  |
|  | Intervention | 11 (32%) |
|  | Prevalence | 9 (26%) |
|  | Other | 5 (15%) |
|  | Diagnostic test accuracy | 3 (9%) |
|  | Risk | 2 (6%) |
|  | Prognosis | 2 (6%) |
|  | Phenomenology | 1 (3%) |
|  | Rate | 1 (3%) |
| **HOW WAS THE STUDY DESIGNED, CONDUCTED AND ANALYSED?** |  |  |
| **What study design did the researchers use?** |  |  |
|  | Cross-sectional | 10 (29%) |
|  | Systematic review | 6 (18%) |
|  | Case series | 4 (12%) |
|  | Scoping review | 2 (6%) |
|  | Mathematical modelling | 2 (6%) |
|  | Cohort with control | 2 (6%) |
|  | Cross over randomised control trial | 1 (3%) |
|  | Descriptive qualitative study | 1 (3%) |
|  | In-vitro | 1 (3%) |
|  | Cohort without control | 1 (3%) |
|  | Narrative or literature review | 1 (3%) |
|  | Clinical practice guideline | 1 (3%) |
|  | Case control | 1 (3%) |
|  | Preclinical animal study | 1 (3%) |
| **Did the method stated match what was described?** |  |  |
|  | Yes | 20 (59%) |
|  | No method given | 13 (38%) |
|  | No | 1 (3%) |
| **Was there a sample size calculation or was the power of the study to provide a meaningful result discussed?** |  |  |
|  | N/A | 21 (62%) |
|  | No | 11 (32%) |
|  | Yes | 2 (6%) |

### Supplementary Figures

**Supplementary Figure 1:**
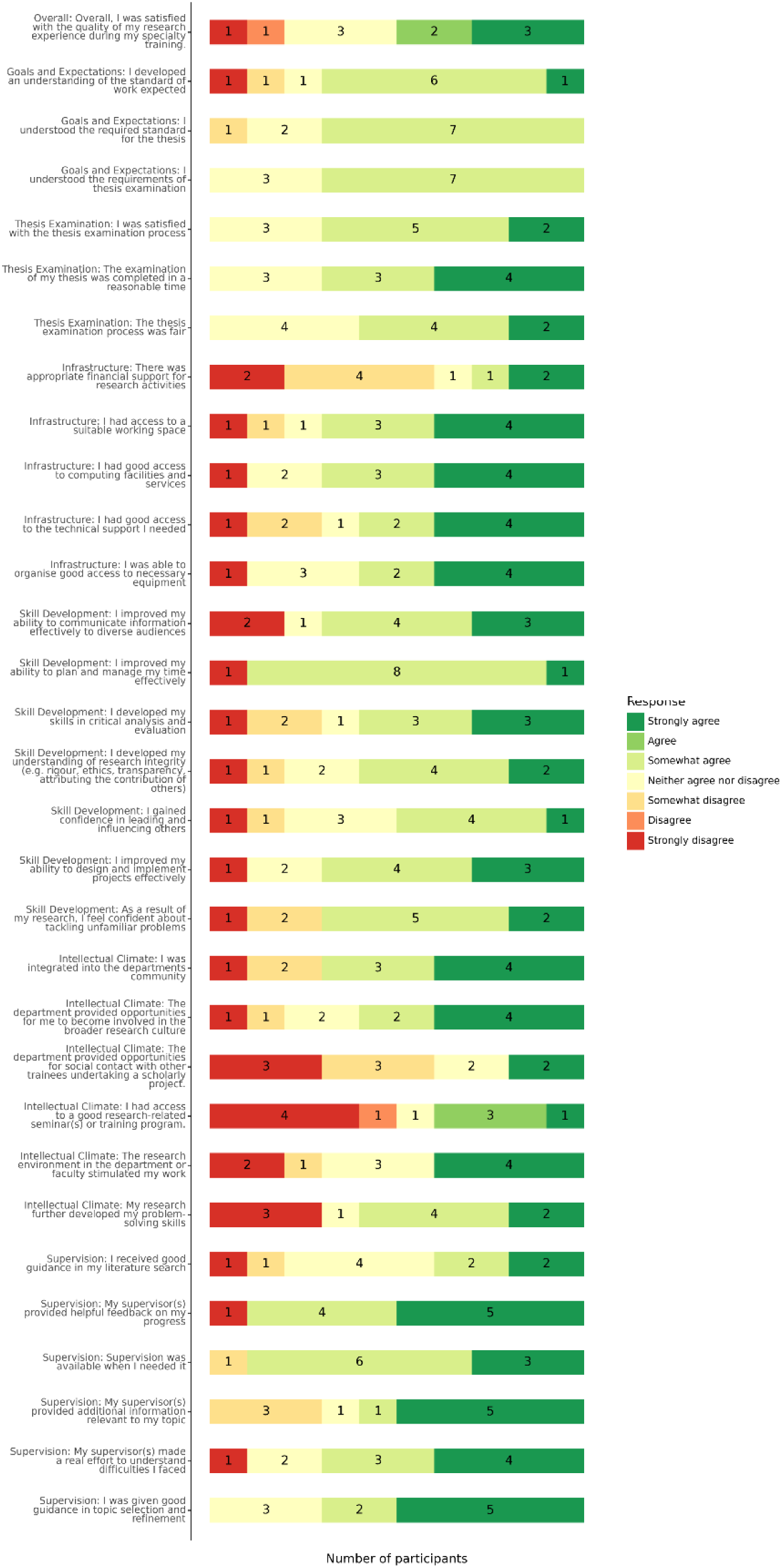
PREQ. Of the 177 eligible participants, the ten respondents completed these sections (6% of eligible participants). PREQ aimed to gain an understanding of trainee satisfaction with the research experience and supervision and indicated the intellectual climate domain and skills development domain scored the worst, whereas the thesis examination and goals and expectations scored the best.

**Supplementary Figure 2:**
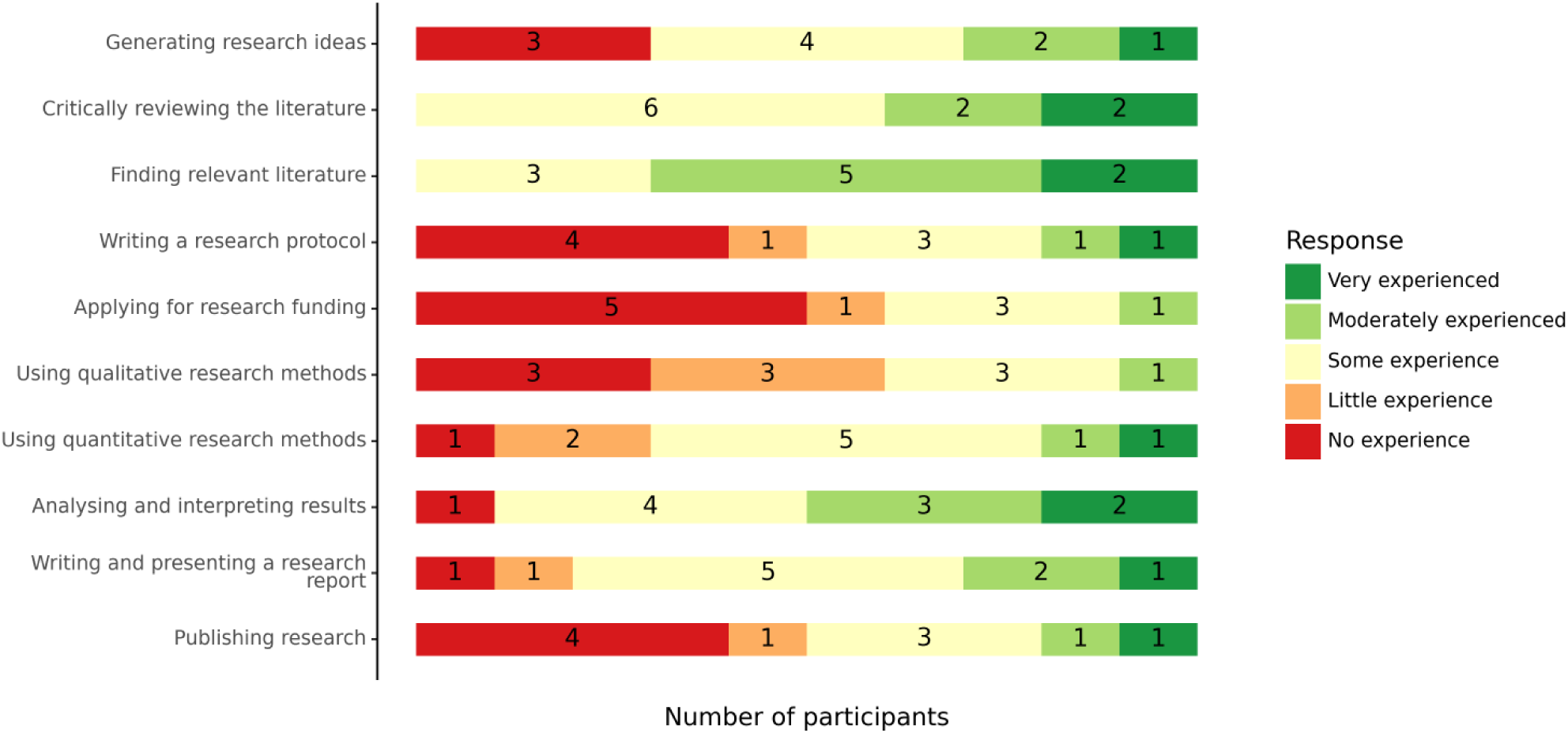
The WReN Spider instrument. This measured trainees’ self-perceived end-of-training research knowledge, skills and experience. Participants felt that, at the end of their research project, they were somewhat to very experienced in finding relevant literature and critically reviewing it. On the other hand, most respondents felt they had no to little experience in writing a research protocol, publishing research, applying for funding, and using qualitative research methods.

**Supplementary Figure 3:**
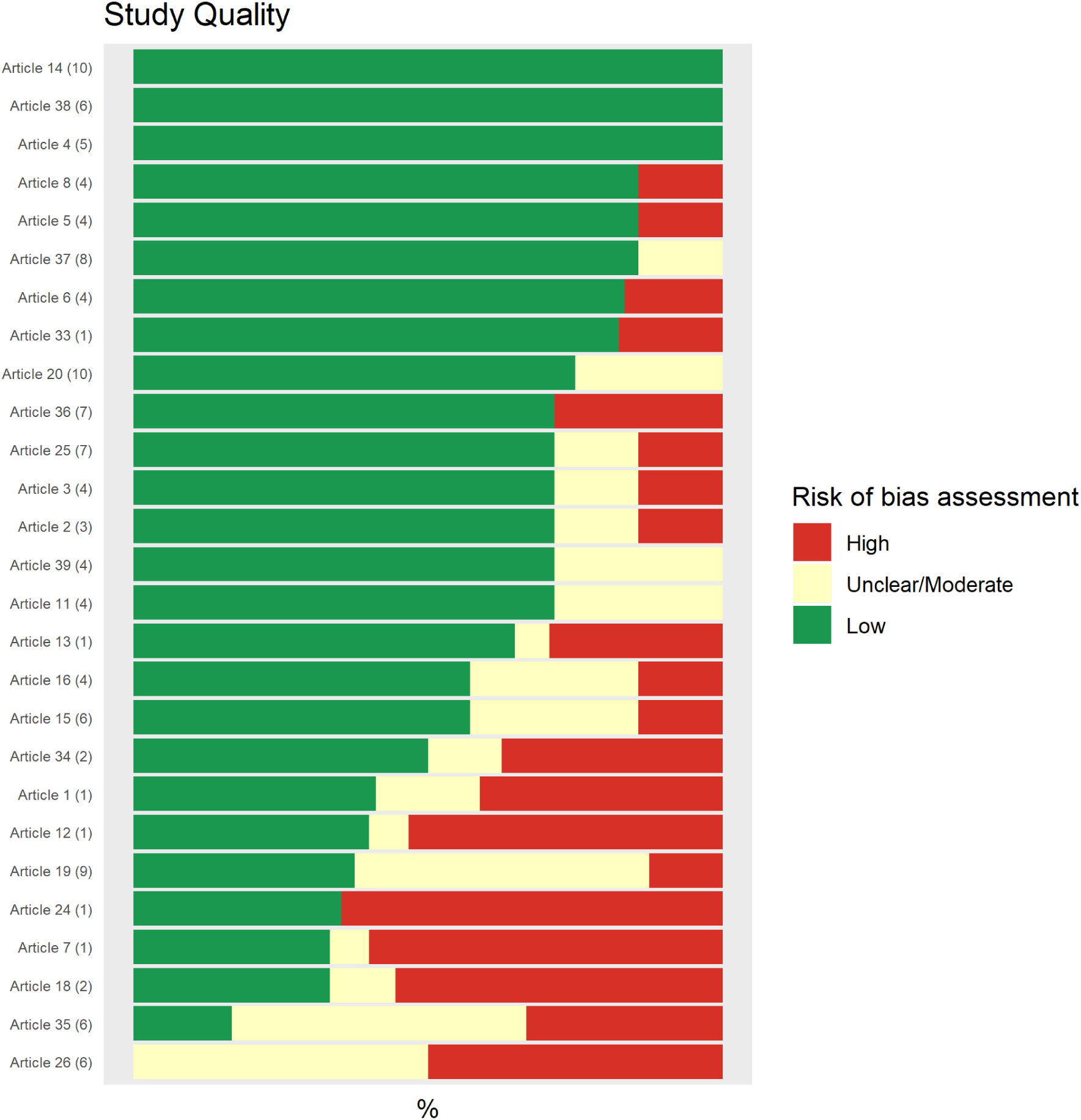
Study Quality. Representation of the study quality of 27 uploaded studies. Numbers in brackets represent the study design. The following critical appraisal tools were used for the following study designs: Modified AMSTAR 2: (1) Review - Systematic Review (2) Review - Scoping Review; MMAT: (3) Randomised Control Trial, (4) Cross sectional, (5) Qualitative, (6) Case series, (7) Cohort WITH control, (8) Randomised preclinical animal study; Mi-Che: (9) Clinical Practice Guideline; QADAS 2: (10) Diagnostic test accuracy study.

